# Circulatory dietary and gut-derived metabolites predict preclinical Alzheimer’s disease

**DOI:** 10.1101/2024.05.10.24307050

**Authors:** Emily Connell, Saber Sami, Mizanur Khondoker, Anne-Marie Minihane, Matthew G. Pontifex, Michael Müller, Simon McArthur, Gwenaelle Le Gall, David Vauzour

## Abstract

A key component of disease prevention is the identification of at-risk individuals. Microbial dysbiosis and microbe-derived metabolites (MDM) can influence the central nervous system, but their role in disease progression and as prognostic indicators is unknown. To identify preclinical factors associated with Alzheimer’s disease (AD), we compared gut microbiome and metabolome profiles of cognitively healthy subjects, subjective cognitive impairment (SCI) participants and mild cognitive impairment (MCI) participants (n=50 per group, matched for age, BMI and sex), targeting metabolites previously associated with cognitive health (TMAO, bile acids, tryptophan, *p*-cresol and their derivatives). 16S rRNA bacterial microbiome sequencing and targeted LC-MS/MS were employed for faecal microbiome speciation and serum MDM quantification. Microbiome beta diversity differed between healthy controls and SCI participants. Multiple linear regression modelling highlighted five serum metabolites (indoxyl sulfate, choline, 5-hydroxyindole acetic acid, indole-3-propionic acid (IPA) and kynurenic acid) significantly altered in preclinical AD. Neuroprotective metabolites, including choline, 5-hydroxyindole acetic acid and IPA, exhibited lower concentrations in SCI and MCI in comparison to controls, while the cytotoxic metabolite indoxyl sulfate had higher levels. A Random Forest algorithm with multiclass classification confirmed and extended our results, identifying six metabolites (indoxyl sulfate, choline, 5-hydroxyindole acetic acid, IPA, kynurenic acid, kynurenine) as predictors of early cognitive decline, with an area under the curve of 0.74. In summary, a combined statistical and machine learning approach identified MDM as a novel composite risk factor for the early identification of future dementia risk.

## 1. Introduction

Currently, an estimated 55.2 million people suffer from dementia worldwide, of which Alzheimer’s disease (AD) is the main form ^1^. In the absence of an effective strategy to slow or prevent disease progression, dementia incidence is expected to increase to 152.8 million by 2050. By the time AD is typically diagnosed, substantial neuronal loss will have occurred across multiple brain regions. Identifying molecular precursors and biological risk factors at preclinical disease stages would enable earlier detection and the targeting of regular monitoring and mitigating interventions while prevention is viable.

The contribution of lifestyle factors to cognitive decline and dementia is well documented ^2, 3^. Diet in particular has emerged as a key influencer of brain health and AD development, in part by modulating communication along the microbiota-gut-brain axis. This axis forms a bidirectional communication system comprising neuronal, endocrine, immune and metabolic signalling mechanisms linking the gut and the central nervous system (CNS) ^4^. Gut microbes regulate this communication via the breakdown of dietary compounds into bioactive metabolites. Such microbe-derived metabolites (MDM) subsequently modulate pathways affecting the CNS both directly, by crossing the blood-brain barrier and indirectly, via modulation of peripheral organ function or vagus nerve stimulation ^5^. In the prodromal stages of AD, for example mild cognitive impairment (MCI), the microbiota-gut-brain axis becomes dysregulated (i.e., dysbiosis), a change associated with pathological processes such as neuroinflammation and neural injury, and thought to contribute to accelerating cognitive decline ^6–8^. However, the mechanism(s) underlying these changes, and the role of MDM in this process remains unknown.

Several examples of MDM have been linked to cognitive health ^5^, including trimethylamine N-oxide (TMAO) ^9–12^, bile acids (BAs) ^13–15^, tryptophan ^16–19^, *p*-cresol and its derivatives ^20, 21^. Notably, these same MDM have been further linked to pathological processes known to be associated with AD ^21–28^, including neuroinflammation, synaptic damage and blood-brain barrier disruption, but whether changes in these MDM are drivers or correlates of disease processes requires comprehensive investigation.

Targeted metabolomics presents a powerful tool to comprehensively assess changes in the endogenous metabolome. Here, we present a targeted metabolomics approach employing liquid chromatography-tandem mass spectrometry (LC-MS/MS) to quantify TMAO, BAs, tryptophan and *p*-cresol metabolite profiles in the serum of healthy controls and participants in early cognitive decline. Early cognitive decline comprises individuals with subjective cognitive impairment (SCI) and mild cognitive impairment (MCI), the preclinical stages of AD progression. This study presents, for the first time, the prognostic value of key metabolites in combination and represents one of only a few studies characterising metabolic perturbations in the early stages of cognitive decline, including participants undergoing the earliest preclinical stage of AD, SCI.

## 2. Materials and methods

### 2.1 Study samples

Human serum samples from the baseline measurements of two previously conducted clinical studies were used: (1) the impact of Cranberries On the Microbiome and Brain in healthy Ageing sTudy (COMBAT; NCT03679533) and (2) the Cognitive Ageing, Nutrition and Neurogenesis (CANN; NCT02525198) study. The COMBAT study recruited 60 adults, aged 50-80 years, with no subjective memory complaints as assessed by the Cognitive Change Index (CCI) questionnaire ^29^. The CANN study recruited 259 participants, aged ≥ 50 years, with subjective cognitive impairment (SCI) or mild cognitive impairment (MCI) based on criteria developed by the National Institute of Aging-Alzheimers Association, with no indication of clinical dementia ^30^. Cognitively healthy adults were selected from the COMBAT study as a control group, with all groups (controls, SCI and MCI, n=50 per group) matched for age, BMI and sex as these are key variables known to affect microbiome composition ^31, 32^. Participants with chronic fatigue syndrome, liver disease, diabetes mellitus, or gall bladder abnormalities were excluded.

Cognitive health was assessed using a variety of cognitive tests in both the COMBAT and CANN study. However, only the Trail Making Test (assessing visual processing speed, scanning, mental flexibility, as well as executive function) and the digit span test (assessing verbal short-term and working memory) were used across the COMBAT and CANN studies enabling comparisons. Participants also completed a validated, semi-quantitative Scottish Collaborative Group (SCG) food frequency questionnaire (version 6.6) to assess background diet ^33^. Biochemical analyses of blood glucose, liver function (bilirubin, albumin, aspartate aminotransferase (AST), alanine aminotransferase (ALT) and AST/ALT ratio), kidney function (creatinine) and serum lipid concentrations (total-, LDL-, HDL-cholesterol and triglyceride) were conducted in all participants. The protocols were approved by the UK National Research Ethics Service (NRES) Committee, (Study ID: 14/EE/0189) for CANN and by the University of East Anglia’s Faculty of Medicine and Health Sciences Ethical Review Committee (Reference: 201819–039) and the UK Health Research Authority (IRAS number: 237251) for COMBAT. The participants provided written informed consent to participate.

### 2.2 Microbiome Profiling

Microbiome analysis was performed by 16S rRNA sequencing as previously reported ^34^. In brief, DNA extraction was performed from approximately 50 mg of faecal content using the QIAamp PowerFecal Pro DNA Kit (Qiagen, Manchester, UK) as per the manufacturer’s instructions. DNA quantity was evaluated using a Nanodrop 2000 Spectrophotometer (Fisher Scientific, UK). Quality assessment by agarose gel electrophoresis distinguished the DNA integrity, purity, fragment size and concentration. Illumina NovaSeq 6000 PE250 was used to amplify the V3–V4 hypervariable region. Sequence analysis was carried out using Uparse software (Uparse v7.0.1001) ^35^, incorporating all the effective tags. Sequences sharing a similarity of ≥97% were grouped into the same Operational Taxonomic Unit (OTU). A representative sequence for each OTU was screened for further annotation. A representative OTU sequence was further analysed using the SSUrRNA database of SILVA Database 138 ^36^. OTU abundance data were normalised using a standard sequence number corresponding to the sample with the least sequences. Alpha diversity was assessed using both Chao1 and Shannon H diversity indices, whilst beta diversity was assessed using Bray–Curtis dissimilarity. Statistical significance was determined by Kruskal–Wallis or Permutational Multivariate Analysis of Variance (PERMANOVA). Comparisons at the phylum and genus level were made using classical univariate analysis using Kruskal–Wallis combined with a false discovery rate (FDR) approach used to correct for multiple testing. P-values below 0.05 were considered statistically significant.

### 2.3 Metabolite Profiling

Serum samples were diluted with methanol at a ratio of 1:10 (v/v) and placed on dry ice for 10 min. Samples were then centrifuged (5 min, 16,000x g at room temp), supernatants filtered using a 0.45 µM PTFE syringe filter and evaporated to dryness using a Savant™ SpeedVac™ High-Capacity Concentrator (Thermofisher, UK). Dried samples were resuspended in either 50 µL of methanol with the addition of 15 µL of lithocholic acid-d4 and cholic acid-d4 at 50 µg/mL for the detection of bile acids, 50 µL water with TMA-d9 N-oxide, TMA 13C 15N hydrochloride at 50 µg/mL for the detection of TMAO/TMA/choline or 50 µL water with 15 µL of L-methionine-3, 3, 4, 4 d4 and p-toluenesulfonic acid at 50 µg/mL for the detection of tryptophan and *p*-cresol metabolites respectively. All internal standards were supplied by Thermofisher, UK. Samples were analysed using the Waters Acquity UPLC system and Xevo TQ-S Cronos mass spectrometer with MassLynx 4.1 software. See supplementary methods for full details.

### 2.4 Statistical Analyses

Significant associations of metabolites with cognitive status were identified using multiple linear regression analysis. Covariates known to affect metabolome or microbiome composition, including age, BMI, diet and markers of kidney function (creatinine) and liver function (AST/ALT ratio), were included in the model ^37–41^. Sex was not included as a covariate as all groups had an equal proportion of males and females. Diet was assessed using a validated, semi-quantitative Scottish Collaborative Group (SCG) food frequency questionnaire (version 6.6) ^33^. Participants’ dietary components were grouped (kcal, proteins, fats, carbohydrates, water, alcohol, vitamins and minerals) and analysed using hierarchical clustering via Ward’s linkage method to assemble individuals with similar dietary patterns (Supplementary Figure S1). This clustered participants into low, moderate and high intake of dietary components and was added to the model as a categorical variable, with participants with a moderate intake used as a reference group. Age, BMI, creatinine and AST/ALT ratio were added to the model as continuous variables. Finally, cognitive status (i.e., control, SCI and MCI) was added to the model as a categorical variable. Metabolite concentrations outside ± 2 standard deviations from the mean were excluded as outliers. The assumptions for multiple linear regression analysis including the existence of a linear relationship among the outcome and predictor variable, normality and homoscedasticity were assessed (Supplementary Figure S2). The model tested for significant associations between metabolite and cognitive status, adjusting for the included covariates. All multiple linear regression analyses were performed in R (v3.6.3; R Foundation: A Language and Environment for Statistical Computing).

### 2.5 Machine Learning

A Random Forest (RF) machine learning algorithm was implemented to assess whether metabolites could be predictive of preclinical AD. The RF model was constructed using 100 decision trees and 6 random variables considered at each split. The number of variables considered per split corresponds to the square root of the total number of attributes in the data ^42^ (as 32 variables were considered, this resulted in ∼6 random variables per split). To create a composite panel to predict preclinical AD, metabolites were ranked according to the mean decrease Gini. This highlights the loss in model performance when permuting the predictor values, and can provide more robust results than mean decrease accuracy ^43^. The metabolites with the highest mean decrease Gini score producing the highest AUC values were retained in the model. To compare our model, Naive Bayes and AdaBoost machine learning models were also constructed ^44, 45^. AdaBoost predictions were made by using a weighted average of weak classifiers. Our model contained 50 estimators, a learning rate of 1.00 and a SAMME classification algorithm which updated the base estimator’s weights with classification results. The Naive Bayes method was applied based on applying Bayes’ theorem with the “naive” assumption of conditional independence between every pair of features given the value of the target variable. The dataset for multi-class classification was randomly divided into training and testing, with 75% of the samples allocated to the training set and 25% to the testing set. Models were assessed by the average area under the receiver operator curve (AUC) (plotting the false positive rate against the true positive rate) over all classes (macro-average) as an indication of model performance. All machine learning models were built in Python (Python Software Foundation. Python Language Reference, version 3.8).

## 3. Results

### 3.1 Study population characteristics

A total of 150 individuals were included in the study of which 50 (33.3%) were cognitively healthy, 50 (33.3%) presented with SCI and 50 (33.3%) with MCI. The mean ± SD age of all participants was 65.5 ± 5.7 years, with a mean level of education of 14.6 ± 3.5 years and 54% female (Table 1). Cognitive groups were matched for age, BMI and sex (p= 0.99). Participants in both the COMBAT and CANN study undertook several cognitive assessments at their baseline visit ^29, 30^. Significant differences were found in the Trail Making Test B, digit span backward test and digit span total score between groups (p<0.05). There was a marginal difference between the three groups in the Trail Making Test A (p= 0.09) and no significant difference occurred in the digit span forwards test (p= 0.21). The prevalence of the *APO* LJ*4* was lower in controls (18%) compared to SCI (26%) and MCI (38%) participants.

**Table 1:**
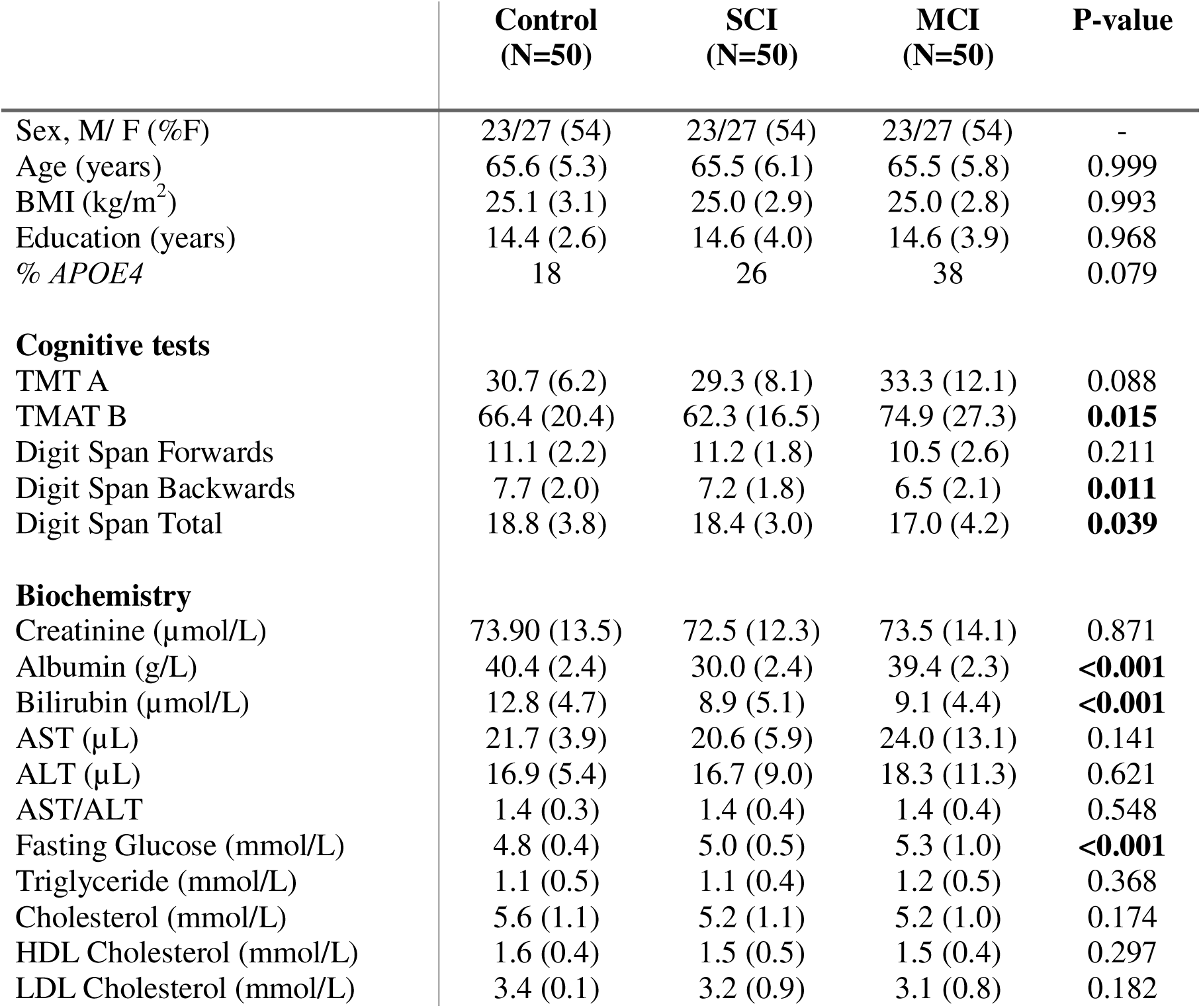
Baseline characteristics of the participants. . Mean (SD). P-value calculated using one-way ANOVA. Significant values at p<0.05 are in bold. SCI: subjective cognitive impairment, MCI: mild cognitive impairment, TMT: Trail Making Test A or B. Bold p-values represent p<0.05.

**Table 2:** Multiple linear regression model (adjusted for age, BMI, liver function (AST/ALT ratio), kidney function (creatinine) and diet) showing metabolites significantly associated with early cognitive decline. Diet was analysed using hierarchical clustering, ‘Ward’ method, to group individuals with similar dietary patterns. This grouped participants into three dietary groups (low, moderate and high intake of dietary components (Kcal, carbohydrates, fats, protein, water, alcohol, minerals, vitamins). Healthy controls and diet group 2 (moderate intake) were used as reference groups in the model. Bold p-values represent *p*<0.05.

| Explanatory Variable | Metabolite |  |  |  |  |  |  |  |  |  |  |  |  |  |  |  |  |  |  |  |
| --- | --- | --- | --- | --- | --- | --- | --- | --- | --- | --- | --- | --- | --- | --- | --- | --- | --- | --- | --- | --- |
|  | Indoxyl Sulfate |  |  |  | Choline |  |  |  | 5-Hydroxyindole Acetic Acid |  |  |  | Indole Propionic Acid |  |  |  | Kynurenic Acid |  |  |  |
|  | Beta | P-value | 95% CI |  | Beta | P-value | 95% CI |  | Beta | P-value | 95% CI |  | Beta | P-value | 95% CI |  | Beta | P-value | 95% CI |  |
|  |  |  | Low | High |  |  | Low | High |  |  | Low | High |  |  | Low | High |  |  | Low | High |
| Constant | -2.288 | 0.482 | -8.705 | 4.128 | 29.690 | 0.011 | 6.938 | 52.443 | 0.068 | 0.020 | 0.011 | 0.125 | 3.094 | 0.009 | 0.790 | 5.397 | 0.017 | 0.448 | -0.027 | 0.062 |
| Age | 0.057 | 0.115 | -0.014 | 0.128 | 0.100 | 0.434 | -0.152 | 0.351 | 0.001 | 0.058 | 0.000 | 0.001 | -0.007 | 0.568 | -0.033 | 0.018 | <0.001 | 0.314 | -0.001 | <0.001 |
| BMI | 0.064 | 0.378 | -0.079 | 0.207 | -0.328 | 0.202 | -0.835 | 0.178 | -0.001 | 0.063 | -0.003 | 0.000 | -0.026 | 0.308 | -0.077 | 0.025 | 0.001 | 0.039 | <0.001 | 0.002 |
| Creatinine | 0.036 | 0.028 | 0.004 | 0.067 | 0.105 | 0.063 | -0.006 | 0.217 | 0.000 | 0.493 | 0.000 | 0.000 | -0.004 | 0.452 | -0.016 | 0.007 | 0.001 | <0.001 | <0.001 | 0.001 |
| AST/ALT | -1.437 | 0.011 | -2.541 | -0.332 | 0.812 | 0.681 | -3.082 | 4.706 | -0.002 | 0.710 | -0.012 | 0.008 | 0.213 | 0.289 | -0.182 | 0.607 | -0.009 | 0.023 | -0.017 | -0.001 |
| Diet Group 1 | 0.354 | 0.445 | -0.559 | 1.267 | 1.247 | 0.445 | -1.972 | 4.466 | -0.002 | 0.690 | -0.010 | 0.006 | 0.204 | 0.221 | -0.124 | 0.532 | -0.003 | 0.408 | -0.009 | 0.004 |
| Diet Group 3 | 0.130 | 0.829 | -1.052 | 1.311 | -0.706 | 0.740 | -4.897 | 3.486 | -0.001 | 0.826 | -0.012 | 0.009 | -0.128 | 0.568 | -0.570 | 0.314 | -0.003 | 0.450 | -0.011 | 0.005 |
| SCI | 1.650 | 0.001 | 0.674 | 2.625 | -5.635 | 0.002 | -9.084 | -2.187 | -0.011 | 0.012 | -0.020 | -0.002 | -0.181 | 0.316 | -0.536 | 0.174 | 0.007 | 0.037 | <0.001 | 0.014 |
| MCI | 1.308 | 0.008 | 0.342 | 2.274 | -5.217 | 0.003 | -8.645 | -1.789 | -0.015 | 0.001 | -0.024 | -0.007 | -0.558 | 0.002 | -0.910 | -0.206 | 0.001 | 0.874 | -0.006 | 0.007 |
| Overall change in early cognitive decline | Increased |  |  |  | Decreased |  |  |  | Decreased |  |  |  | Decreased |  |  |  | Increased |  |  |  |

Albumin, bilirubin and fasting glucose (p<0.01) differed according to cognitive status. Interestingly, both albumin and bilirubin were highest in controls and lowest in SCI participants. Although participants diagnosed with diabetes mellitus were excluded, fasting glucose increased over preclinical AD, with the lowest concentrations in control individuals and the highest in MCI (Table 1).

### 3.2 Gut microbiome and metabolome shifts in preclinical AD

Alpha diversity was measured using the Chao1 (p= 0.21) and Shannon H (p= 0.70) indices with no significant difference amongst groups (Figure 1A-B). Conversely, beta diversity, as measured by Bray-Curtis dissimilarity, was significantly different (PERMANOVA F-value= 1.35, p= 0.02) (Figure 1C). Pairwise analysis suggested the shift was primarily driven by the differences between the control and SCI groups (FDR q= 0.03), rather than between SCI and MCI (FDR q= 0.38) or MCI and control (FDR q= 0.15) (Figure 1D). The PLS-DA plot suggested similar patterns in participants’ metabolomic profiles, with control separating from SCI and MCI (Figure 2A). The extent of this separation can be seen through the heatmap displaying shifts in metabolite concentrations between groups and clustering SCI and MCI together (Figure 2B). The similarity between microbiome and metabolomic profiles was confirmed by conducting a Procrustes analysis to evaluate the congruence of the two datasets. The analysis revealed strong similarity between the metabolome and microbiome results in control and SCI (R= 0.21, p= 0.03), SCI and MCI (R= 0.27, p= 0.002) and MCI and control (R= 0.26, p= 0.002) groups (Supplementary Figure S3).

**Figure 1:**
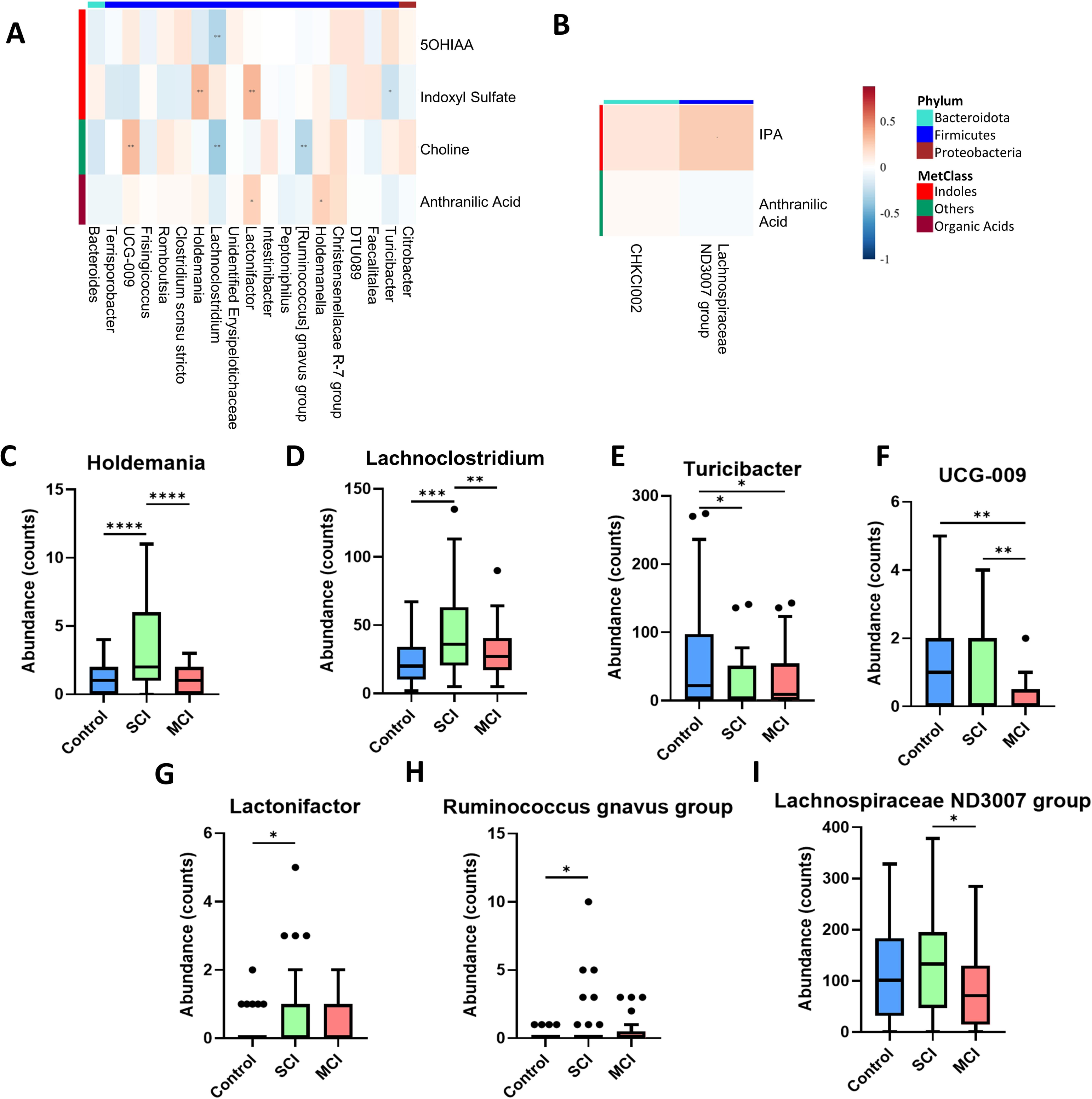
Microbiome beta diversity is significantly altered in early cognitive decline. Alpha diversity measured by Chao1 (A) and Shannon H (B) index. (C) Beta diversity as measured by Bray-curtis; p-value generated from PERMANOVA. (D) Pairwise comparisons of the beta diversity analysis.

**Figure 2:**
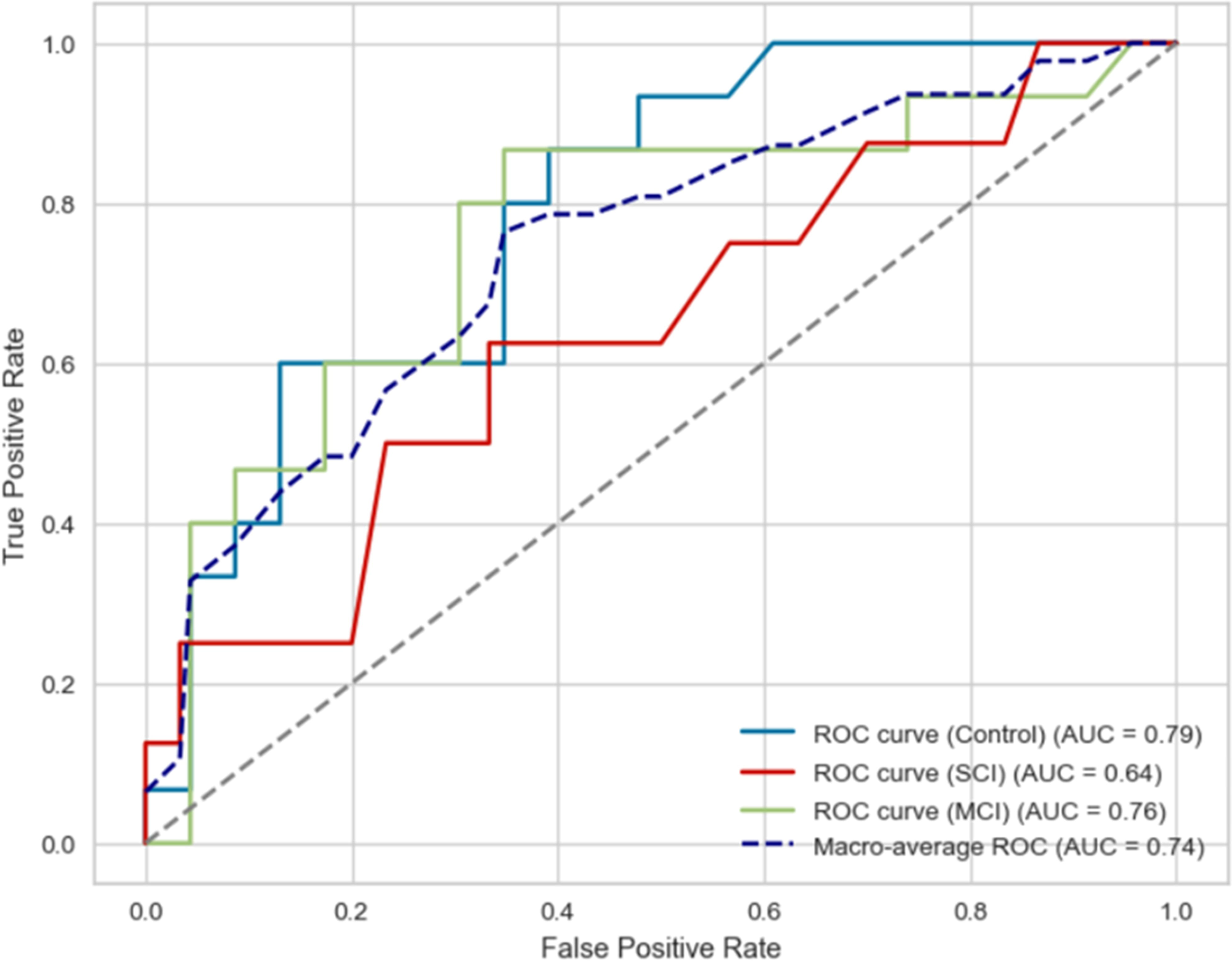
Metabolic shift occurs in early cognitive decline. (A) Partial least squares-discriminant analysis (PLS-DA) plot of the metabolomic profiles. (B) Heatmap displaying changes in concentrations of metabolites between the groups, with hierarchical clustering.

### 3.3 Serum metabolites significantly associated with early cognitive decline in an adjusted multivariable model

Multiple linear regression analysis adjusted for liver function (AST/ALT ratio), kidney function (creatinine), age, BMI and background diet identified five metabolites significantly associated with early cognitive decline, including choline, 5-hydroxyindole acetic acid, indole-3-propionic acid (IPA), indoxyl sulfate and kynurenic acid (Table 1). Indoxyl sulfate, choline and 5-hydroxyindole acetic acid were associated with both SCI and MCI (p<0.05). Kynurenic acid was significantly associated with SCI (β= 0.007, 95% CI: <0.001, 0.014, p= 0.037) but not MCI (β= 0.001, 95% CI: - 0.006, 0.007, p= 0.874). On the other hand, IPA was significantly associated with MCI (β= -0.558, 95% CI: -0.910, -0.206, p= 0.002), but not SCI (β= -0.181, 95% CI: -0.536, 0.174, p= 0.316). Neuroprotective metabolites, including choline, 5-hydroxyindole acetic acid and indole propionic acid ^46–48^, exhibited lower concentrations in SCI and MCI participants in comparison to controls, while metabolites linked to cytotoxicity, including indoxyl sulfate, showed increasing levels ^49^. Kynurenic acid, a typically neuroprotective metabolite ^50^, was higher in SCI and MCI in comparison to controls. Group means for all metabolite concentrations are given in Supplementary Table S1.

### 3.4 Machine learning models to identify risk factors predictive of preclinical AD

All 32 serum metabolites were initially evaluated as possible predictors of preclinical AD. RF achieved the highest classification AUC of 0.65, with AdaBoost and Naïve Bayes attaining 0.58 and 0.63 respectively (Supplementary Table S2). Using the mean decrease Gini, the importance of each metabolite was assessed (Figure S4). Six metabolites (5-hydroxyindole acetic acid, indole-3-propionic acid, choline, indoxyl sulfate, kynurenic acid and kynurenine) produced the highest AUC of 0.74 using the RF classification algorithm (Figure 4). In comparison, Naive Bayes achieved an AUC of 0.72 and AdaBoost attained 0.68. The RF ROC curve indicated the model’s predictive performance was highest for controls (AUC= 0.79), followed by MCI (AUC= 0.76) and SCI (AUC= 0.64). As such, we investigated whether the model performance would be improved by predicting only healthy ageing and MCI. Using the six serum metabolites from controls and MCI participants, the RF model showed improved predictive performance (AUC= 0.84) (Supplementary Table S2). AdaBoost and Naive Bayes also demonstrated increased performance, attaining AUC of 0.87 and 0.90 respectively.

### 3.5 Serum dietary and MDM and gut microbiome modulation in preclinical AD are significantly linked

Having identified a shift in both the microbiome and metabolome profiles, a Spearman correlation examined possible connections between the two datasets. Significantly modulated metabolites and microbiome genera between the three groups (p<0.05) were correlated, revealing bacterial–metabolite interactions. Control and SCI participants displayed a negative relationship between 5-hydroxyindole acetic acid and *Lachnoclostridium* (R= -0.29, p=0.004), indoxyl sulfate and *Turicibacter* (R= -0.21, p= 0.038), and choline and *Lachnoclostridium* (R= -0.36, p= 0.0002) and *Ruminococcus gnavus* group (R= -0.28, p= 0.005) (Figure 3A). Choline and *UCG-009* (R= 0.316, p= 0.002), anthranilic acid and *Lactonifactor* (R= 0.24, p= 0.019) and *Holdemanella* (R= 0.24, p= 0.02), indoxyl sulfate and *Holdemania* (R= 0.318, p= 0.001) and *Lactonifactor* (R= 0.33, p=0.0008) had a positive correlation. Between SCI and MCI participants, only IPA and *Lachnospiraceae ND3007* group had a positive correlation (R= 0.26, p= 0.011) (Figure 3B).

**Figure 3:**
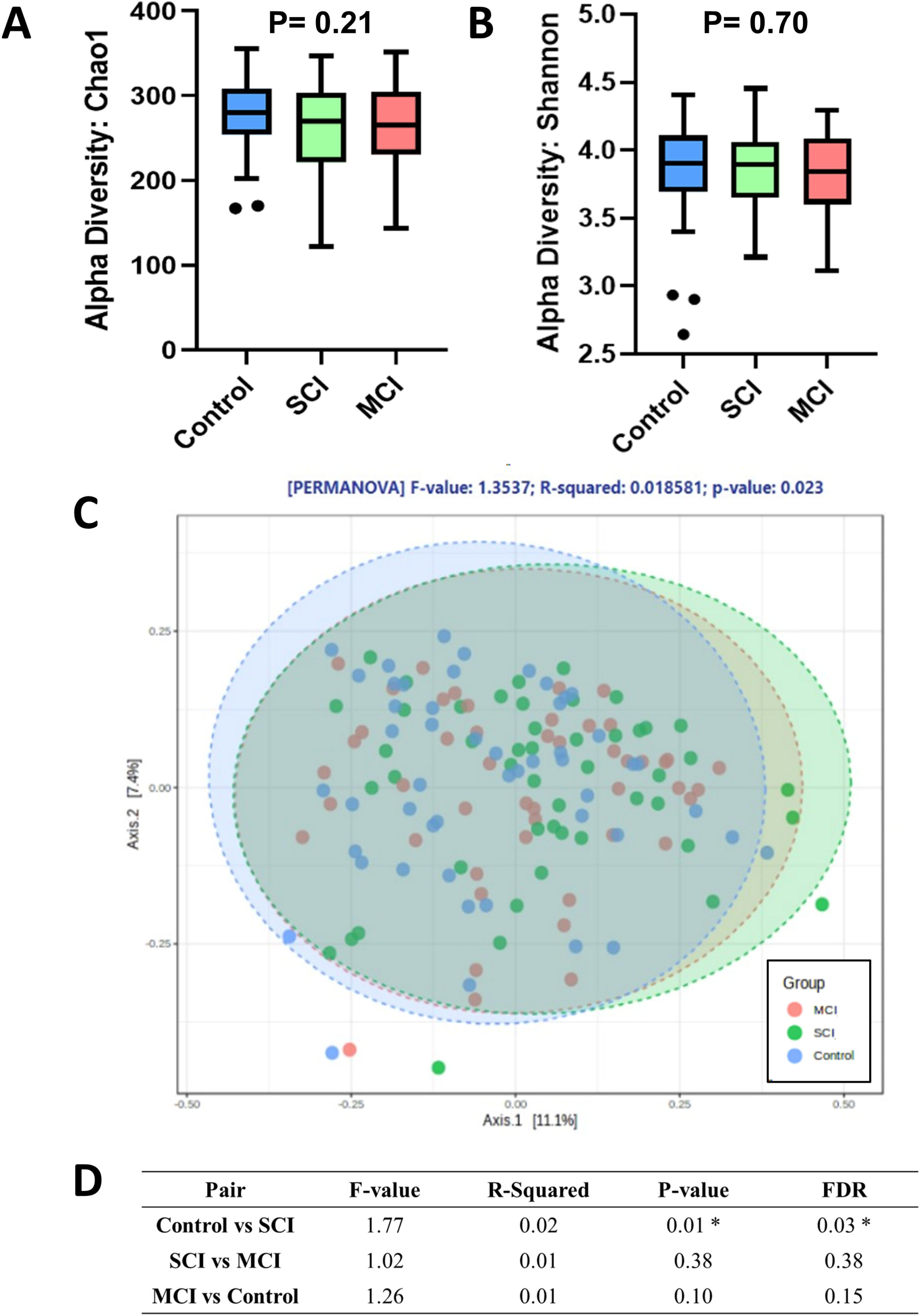
Serum metabolome and gut microbiome profiles are linked. Spearman rank correlation analysis between metabolite and microbiome genera that are significantly modulated in early cognitive decline (A) between control and SCI and (B) between SCI and MCI. (C-H) Abundance counts of microbiome genera correlated with our metabolites of interest (indoxyl sulfate, choline and 5-hydroxyindole acetic acid) between control and SCI participants. (I) Abundance count of microbiome genera correlated with metabolite of interest (indole-3-propionic acid) between SCI and MCI, *=p<0.05, **=p<0.01.

**Figure 4:**
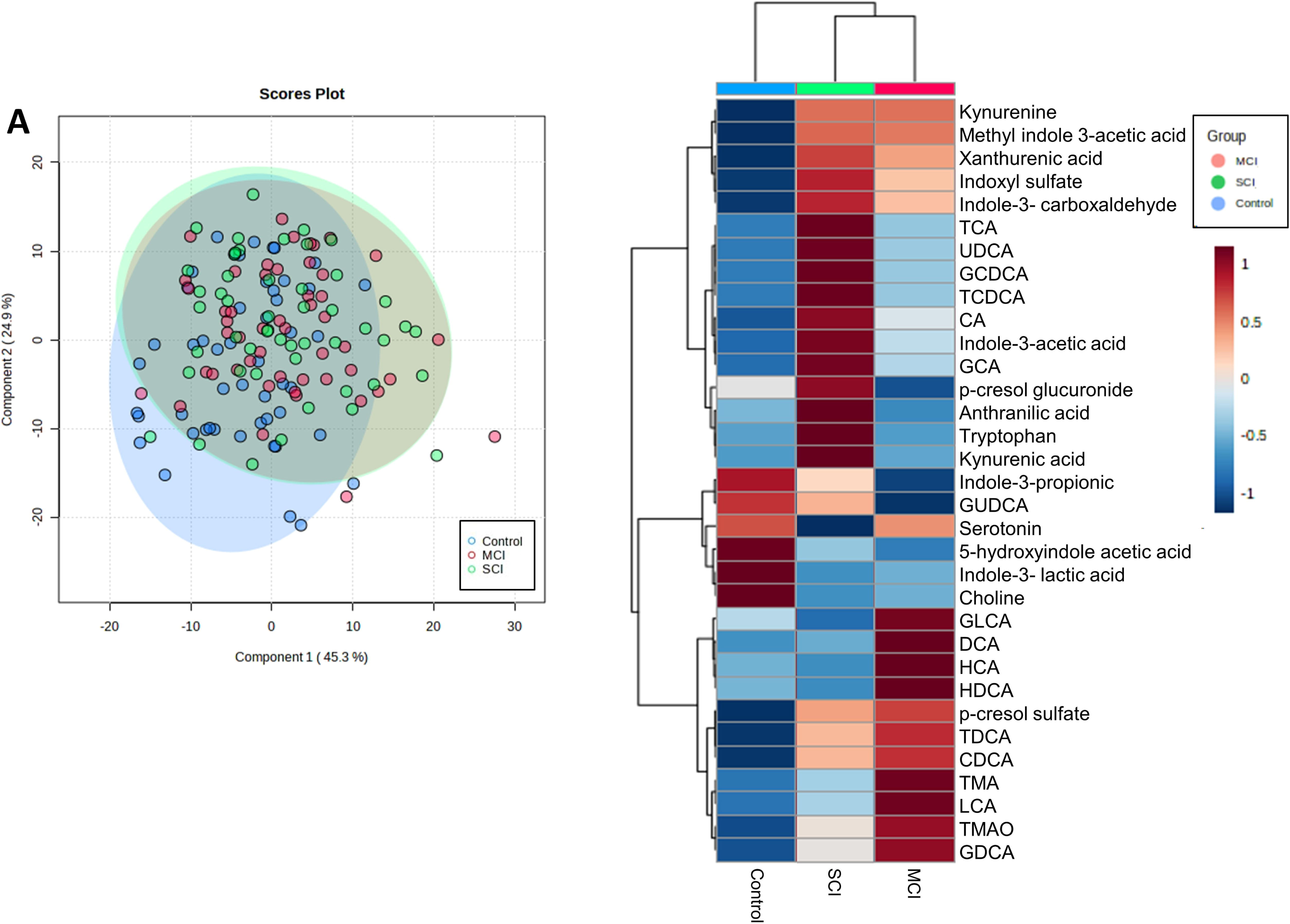
Six circulatory metabolites are predictive of preclinical AD. Receiving Operating Characteristic (ROC) curve illustrating the performance of the Random Forest model for classifying controls, SCI and MCI participants with average area under the curve (AUC) of the multilevel classifier.

As indoxyl sulfate, choline, 5-hydroxyindole acetic acid and IPA were identified as risk factors of cognitive decline, genera correlated with these metabolites were investigated further to identify potential associations with their production. *Holdemania* and *Lachnoclostridium* genera were higher in SCI participants in comparison to controls but lower in SCI relative to MCI (Figure 3C-D). On the other hand, *Turicibacter* were higher in both SCI and MCI participants in comparison to controls (Figure 3E). The abundance of *UCG-009* was significantly lower in both SCI and MCI participants in comparison to controls (Figure 3F). *Lactonifactor* and *Ruminococcus gnavus* were both higher in SCI in comparison to controls (Figure G-H). Finally, *Lachnospiraceae ND3007* was not different between control and SCI but was lower in SCI compared to MCI (Figure 3I).

## 4. Discussion

Identification of robust, inexpensive and non-invasive markers of cognitive status and its trajectory is currently an unmet medical need in AD research, with circulating gut-derived metabolites presenting a promising area. Metabolic alterations contain rich systemic information on the underlying physiology that connects the periphery to the CNS, likely affecting numerous pathways simultaneously. Thus, the simultaneous detection of numerous perturbed metabolites can provide a powerful detection tool. However, studies investigating composite markers are lacking.

16s rRNA sequencing indicated that significant shifts in gut microbiome composition occur during preclinical AD, commencing as early as SCI, suggesting changes may already be apparent when memory complaints first appear, aligning with previous studies ^51, 52^. As cognitive decline progresses from SCI to MCI, gut microbiome modulation appears to be less significant. Circulatory metabolites also reflect this pattern, clustering SCI and MCI participants independently from the healthy controls. Individuals with SCI are likely at a higher risk of cognitive decline progression compared to those who are cognitively healthy ^53^, which may lead to greater alterations in biological markers, including the gut microbiome and its metabolites. Procrustes analysis showed significant congruence of the microbiome and metabolome datasets, suggesting the two are interlinked and provides the potential for MDM predictors to be detected early in disease progression. Indeed, gut microbiome composition can account for up to 58% of the variation of circulatory metabolites communicating along the microbiota-gut-brain axis ^54^.

Targeted metabolomics quantifies metabolites with extremely high sensitivity and accuracy, providing an advantage over the relative responses yielded by untargeted approaches. RF and multiple linear regression models both revealed indoxyl sulfate, choline, 5-hydroxyindole acetic acid, IPA and kynurenic acid as key early indicators of cognitive decline, with RF presenting an AUC predictive performance of 0.74, strongly supporting a significant link between metabolic perturbations associated with the gut microbiome and preclinical AD progression. Previous studies have predominantly concentrated on binary classification approaches, primarily utilising MRI and PET imaging modalities, to investigate AD progression ^55–57^. However, in clinical practice, multiclass classification of blood samples of patients with SCI, MCI and healthy controls could provide a useful approach. Tong and colleagues attained a similar predictive performance (AUC= 0.729) using RF and nonlinear graph fusion of multiple modalities (regional MRI volumes, voxel-based FDG-PET signal intensities, CSF biomarker measures and genetic information) to classify control, MCI and AD participants ^58^. AUC increased to 0.84 when predicting healthy ageing and MCI, likely due to the difficulty of diagnosing a patient undergoing SCI. Indeed, Purser and colleagues found no relationship between memory complaints and the progression of cognitive impairment over 10 years in individuals 65 years and over ^59^. However, others dispute this result ^60^. Adjusting our statistical analysis for confounding variables that heavily influence the host, such as age, BMI, kidney function, liver function and background diet, improves analysis robustness and sensitivity. Adjusting for background diet becomes particularly vital when examining MDM as the diet can both modulate gut microbiome composition and provide a variety of bioactive precursor compounds; a factor which is often overlooked in metabolomic analyses ^61^. Nevertheless, our results highlight the use of profiling select circulatory MDM to identify higher-risk individuals of cognitive decline.

Of the five metabolites highlighted by both machine learning and multiple linear regression, all except choline are produced from tryptophan metabolism, indicating notable alterations in tryptophan metabolism may occur in preclinical AD progression. Tryptophan metabolism has previously been well-linked to AD ^18^. Indeed, we find lower neuroprotective tryptophan-derived metabolites, including IPA and 5-hydroxyindole acetic acid, as cognitive decline progresses from controls to SCI and MCI. IPA is produced in the gut by the microbial conversion of tryptophan via the indole pathway and has previously been investigated as a possible treatment for AD ^62^ due to its potent antioxidant effect against Aβ 1-42 *in vitro* ^63^ and its ability to prevent aggregation and deposition of Aβ monomers ^64^. IPA is anti-inflammatory, reducing the concentration of the proinflammatory TNF-α in activated microglia ^65^, lowering the expression of chemokine (CC Motif) ligand 2 (CCL2) and nitric oxide synthase 2 (NOS2) in interferon-beta (IFN-β) activated murine astrocytes ^66^ and preventing increases in cytokines in LPS-induced human primary astrocytes ^67^, and has previously been identified as a predictor of AD progression ^68^. 5-hydroxyindole acetic acid is often used as a surrogate marker for serotonin due to serotonin’s rapid degradation. As such, our findings indicate lower peripheral serotonin breakdown as early cognitive decline progresses. Approximately 95% of all serotonin is localised in peripheral compartments where it is involved in the modulation of the enteric nervous system (ENS) development and neurogenesis, gut motility, secretion, inflammation, and epithelial development, suggesting these processes may be disrupted in early cognitive decline ^69^. Indeed, MCI and AD patients have often been reported to suffer from gastrointestinal symptoms ^70^ and ENS dysregulation in AD has previously been described ^71^. Decreased concentrations of 5-hydroxyindole acetic acid also suggest a shift in tryptophan metabolism towards the kynurenine pathway, reducing the availability of tryptophan for serotonin synthesis. This is supported by higher serum kynurenine concentrations in SCI and MCI participants in comparison to controls (Supplementary Table S1) and has previously been found in AD participants, linked to poor memory, executive function and global cognition ^72^. The kynurenine pathway is activated by an inflammatory stimulus, promoting indoleamine 2,3-dioxygenase, the rate-limiting enzyme that initiates the kynurenine pathway. Increased inflammation is a common feature of AD and as such may play a role in modulating tryptophan catabolites.

Both indoxyl sulfate and kynurenic acid concentrations were increased as cognitive decline progressed, even after adjusting for measures of liver and kidney function. As a uremic toxin, indoxyl sulfate can disrupt neuronal efflux transport systems, promote the production of free radicals, inflammation, endothelial cell dysfunction and disturb the circadian rhythm involved in clearing renal and CNS toxins ^73, 74^, likely contributing to cognitive decline. Serum levels of indoxyl sulfate, as well as albumin, have previously been identified as predictive of cognitive impairment in participants with end-stage renal disease ^75^. End-stage renal disease patients have also been reported to have an increased abundance of the gut bacteria *Holdemania*, in line with our results, suggesting his genera may be underlying the changes between control and SCI ^76^. Rodent studies show increased kynurenic acid concentrations can impair cognitive function, including spatial working memory, and broad monitoring deficits ^77, 78^. However, data regarding this relationship in human studies is inconsistent ^79, 80^. Kynurenic acid can play a protective role against the cytotoxic product of the kynurenine pathway, quinolinic acid, by acting as an NMDA antagonist for both glycine and glutamate modulatory sites ^81^. However, abnormal accumulation has previously been found to induce glutamatergic hypofunction and subsequently disrupt cognitive function ^82^. In AD, increased blood concentrations of kynurenic acid have been hypothesised to relate to neuroinflammatory processes and may be produced as a protective response to neuronal damage ^83^.

Choline is required for numerous biological functions in the body ^84^, notably including hallmark AD-associated processes such as acetylcholine synthesis ^85^. As choline readily crosses the blood-brain barrier, peripheral concentrations typically mirror concentrations in the brain ^86^, thus lower concentrations in early cognitive decline may indicate decreased central acetylcholine production. Acetylcholine is intricately connected to neural networks regulating memory, and a reduction in this system is closely associated with learning and memory deficits in AD ^87^. *Lachnoclostridium* and *Lactonifactor* were inversely correlated with choline levels, suggesting changes in these genera may modulate blood concentrations. Indeed, previous research has found *L. saccharolyticum* WM1, a representative strain of *Lachnoclostridium*, to be an efficient converter of choline to TMA *in vitro*, transforming at a rate near 100% ^88^. This metabolic process *in vivo* also elevated serum TMAO levels ^88^, which is supported by our results displaying a 1.6-fold higher TMAO in MCI compared to controls. It is likely that increases in *Lachnoclostridium* abundance may increase the metabolism of choline to TMAO, decreasing its concentration in circulation.

Our study has major strengths including simultaneously targeting some of the top microbial and metabolic metabolites associated with cognitive decline, whilst matching our participants and adjusting our analysis for key factors known to influence the metabolome (age, BMI, sex, liver function, kidney function and background diet), factors rarely accounted for in marker studies. Furthermore, our study highlights key microbiota underlying these metabolic changes, as well as investigating participants from the earliest stage of decline (SCI) and validating our results through machine learning and adjusted statistical approaches. However, some limitations should be stressed. Despite our study adjusting results for key covariates, host metabolome profiles are influenced by a plethora of additional largely environmental and biological factors. Thus, although our findings suggest relationships between the variables, we cannot infer causal relationships from this analysis alone. Moreover, participants’ background diet was adjusted for using data collected by food frequency questionnaires, which can be prone to measurement error and may introduce inaccuracies due to recall bias and self-reporting issues. Furthermore, like all studies utilising machine learning models, the larger the dataset, the more robust the predictive performance. With the current dataset including 150 individuals and 32 metabolites, we achieved significant predictive performance. However, our findings will require external validation in larger independent cohorts to improve the model.

Pathophysiological progression of AD is apparent up to 20 years prior to clinical symptom onset, making it vital for prevention research to focus on uncovering novel preclinical risk factors. Scalable markers that enable the early detection of at-risk persons could permit the targeting of lifestyle interventions to lessen future risk and uncover novel mechanisms underpinning dementia. Our findings present new insights into the preclinical progression of cognitive decline and dementia. We signify a major role for the gut in connection to the brain through the modulation of key MDM. Furthermore, we lend strength to the hypothesis that individuals with higher risks of cognitive decline can be identified via a targeted metabolomic approach in the preceding stages of AD.

## Data availability

The 16S rRNA gene sequence data have been deposited in the NCBI BioProject database (https://www.ncbi.nlm.nih.gov/bioproject/) under accession number PRJNA1109848. Other data that support the findings of this study are available from the corresponding authors upon reasonable request.

## Authors Contributions

D.V. conceptualised the project. A.M.M. provided the human blood and faecal samples. G.L.G. designed the metabolomic analysis, which E.C. and G.L.G. then performed. E.C. conducted the 16S rRNA microbiome sequencing. E.C. and S.S. performed the machine learning analysis. E.C. and M.K. conducted the statistical analysis. E.C. wrote the paper. D.V., S.S., M.K., A.M.M., M.G.P., M.M., S.M. G.L.G. contributed key discussions in interpreting results and edited the paper. All authors approved the final version of the paper.

