## Supplementary Tables and Figures for "Circulatory dietary and gut-derived metabolites predict preclinical Alzheimer’s disease"

**1. Supplementary Methods of LC-MS/MS**

1.1 Sample collection

Overnight fasted blood samples were drawn from participants during their baseline study visit of the COMBAT and CANN studies. After collection, blood was left to coagulate (in clot-activating gel tubes), followed by centrifugation at 2,000g for 10 min and removal of the resultant serum from the sub-fractions. Aliquoted serum was stored at −80°C until further analysis.

1.2 Sample preparation

Serum samples were diluted with ice-cold methanol at a ratio of 1:10 (*v/v*) and placed on dry ice for 10 min. Samples were centrifuged (5 min, 16,000x g at room temp), supernatants filtered using a 0.45 µM PTFE syringe filter and evaporated to dryness using a Savant™ SpeedVac™ High-Capacity Concentrator (Cat. SC210A-230). For the detection of bile acids, dried samples were resuspended in 50 µL of methanol with 15 µL of lithocholic acid-d4, cholic acid-d4 at 50 µg/mL as the internal standards. For the detection of TMAO/TMA/choline, dried samples were resuspended in 50 µL water with TMA-d9 N-oxide, ^13^C_3_^15^N TMA hydrochloride at 50 µg/mL as the internal standards. Finally, for the detection of tryptophan and *p-cresol-related* metabolites, dried samples were resuspended in 50 µL water with 15 µL of L-methionine-3, 3, 4, 4 d4 and p-toluenesulfonic acid at 50 µg/mL as the internal standards for tryptophan and *p*-cresol metabolites respectively.

Stock solutions of each metabolite were prepared in methanol (1mg/mL) and stored at -80°C. Calibration standards were prepared by pooling all relevant analytes for each method at eight concentrations and adding the respective internal standards at 50 µg/mL. Calibration standards were run at the beginning, middle and end of each analytical queue. The analyte: internal standard response ratio was used to create calibration curves and quantify each metabolite.

1.3 LC-MS/MS Condition

Metabolite quantification was performed using liquid chromatography-tandem mass spectrometry (LC-MS/MS) comprising of Waters Acquity UPLC system and Xevo TQ-S Cronos mass spectrometer controlled by MassLynx 4.1 software. For the detection of bile acids, the electrospray ionisation (ESI) operated in negative mode and chromatographic separations were performed with a Supelco Ascentis Express C18 column (150 x 4.6 mm, 2.7 µM) (adapted from ^1^). Eluent A (10mM ammonium acetate, 0.1% formic acid, water) and eluent B (10mM ammonium acetate, 0.1% formic acid, methanol ran at a constant rate of 0.6 mL/min. The gradient began at 50% B and was held for 2 min before a linear increase to 95% B occurred at 20 min. This was held for 4 min before a linear decrease in gradient back to 50% B occurred between 24 and 25 min. The gradient was held at 50%B for another 4 mins.

For the detection of TMAO, TMA and choline, ESI operated in positive mode (adapted from ^2,3^). Chromatic separation occurred using a BEH Amide (150 x 2.1 mm, 1.7 µM) and 0.5mL/min using eluent A (10mM ammonium formate, 0.2% formic acid, 50% acetonitrile) and eluent B (10mM ammonium formate, 0.2% formic acid, 95% acetonitrile) initially at 100% B to 60% B at 4 min and held until 4.2 min, before increasing back to initial conditions of 100% B at 4.21 min and being held until 5.4 min for equilibration.

ESI operated in positive mode for the detection of tryptophan and *p-cresol-related* metabolites and chromatic separation occurred using an ACQUITY UPLC BEH C18 1.7 µM (2.1 x 50mm) column at a rate of 0.3 mL/min and composition of eluent A (0.1% formic acid, water) and eluent B (0.1% formic acid, methanol) at a gradient of 5% B from 0 to 0.5 min, 10% B from 0.5 to 2.5 min, 15% B at 3.5 min, 35% B at 4.5 min, 45% B at 6.5 min, 55% B at 7 min, 100% B at 7.5 min, 100% B until 10 min, and 5% B at10.1 min to return to initial conditions for equilibration until 14 mins. This method was adapted from Anesi and colleagues ^2^. Chromatogram peak analysis was performed by the accompanying Waters® TargetLynx ™ application manager and all further data analysis and calibration curve constructions were completed in Microsoft Excel (2019 version).

1.4 Method Performance

Excellent linear response range was ensured for each calibration curve with correlation coefficients (r^2^) 0.99 or higher for all calibration curves generated. An agilent high-performance autosampler with an injection program was used to minimise carry-over effects between samples. Samples were run in a random order and 20% of the whole set was re-run as a quality control for the method repeatability. No signal was also detected in blank samples run amongst the serum and calibration injections, or in blanks run after the highest calibration standard, indicating that there was little to no carry-over occurring.

**Supplementary Figures**

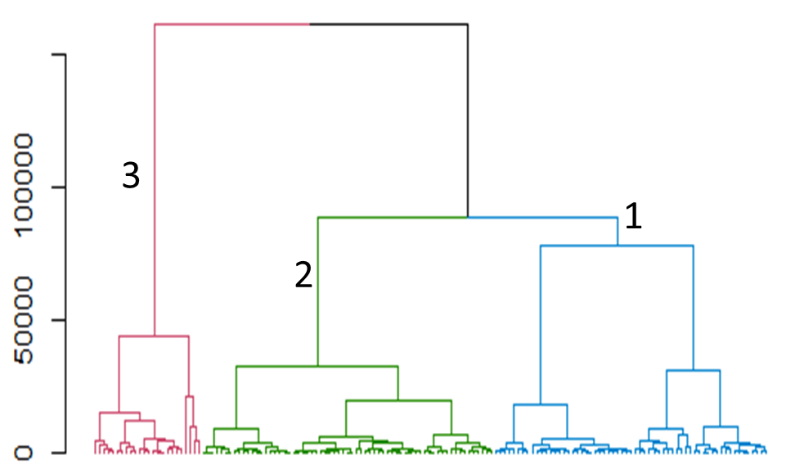

**Figure S1: Hierarchical clustering of participant food frequency questionnaires.** Food frequency questionnaires were analysed using the hierarchical clustering ‘Ward’ method to cluster participants with similar dietary patterns. This grouped participants into low intake of macronutrients (1), moderate intake of macronutrients (2) and high intake of macronutrients (3).

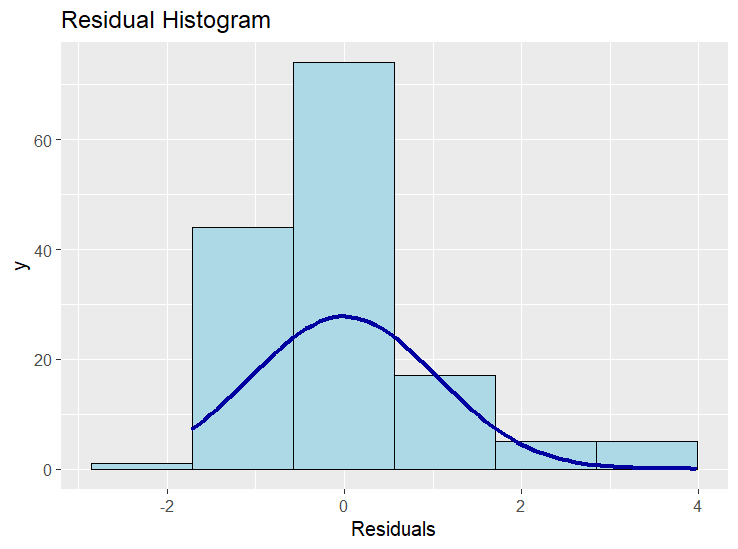

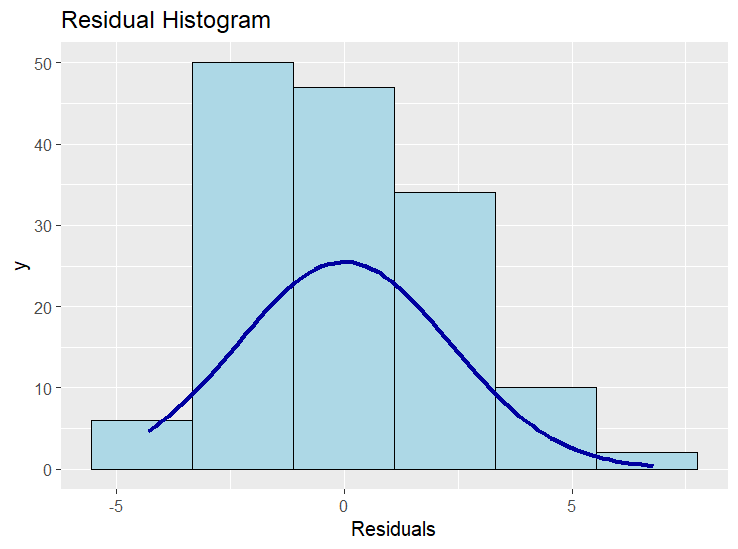

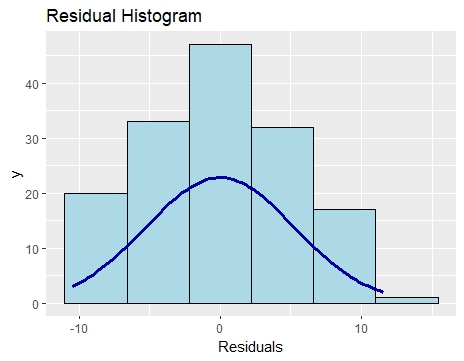

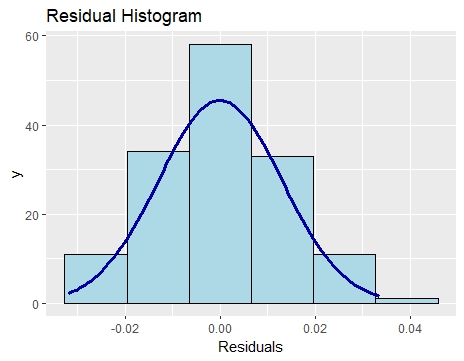

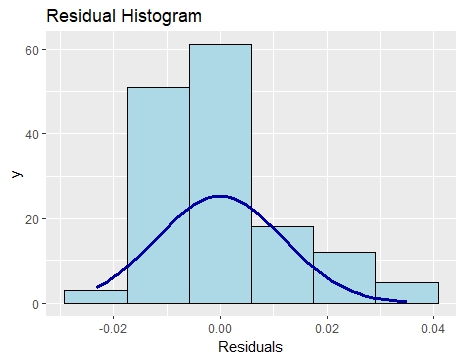

C

B

A

E

D

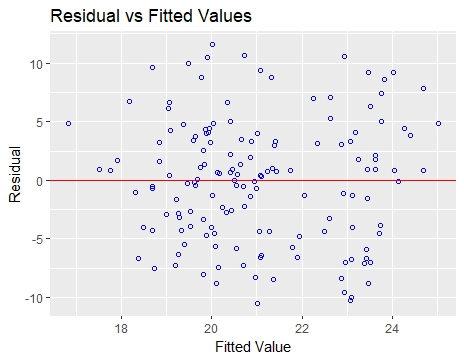

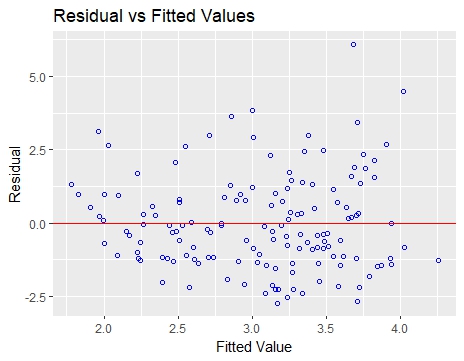

F

G

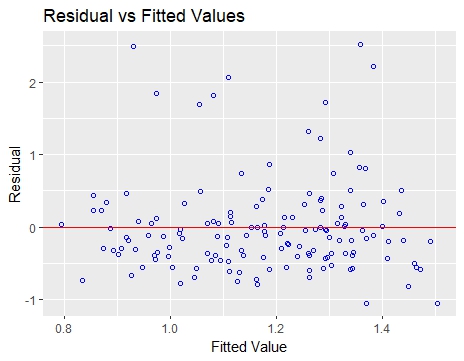

H

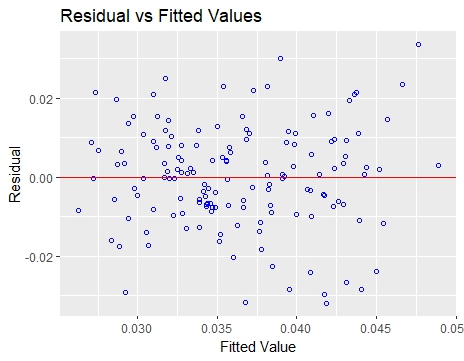

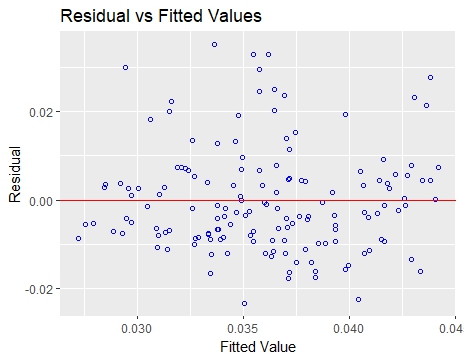

J

I

**Figure S2: Assumptions of the multiple linear regression analysis**. Histogram displaying normality of the residuals for (A) choline, (B) indoxyl sulfate, (C) indole propionic acid, (D) kynurenic acid, (E) 5-hydroxyindole acetic acid. Residuals versus fitted plot confirming homoscedasticity of (F) choline, (G) indoxyl sulfate, (H) indole propionic acid, (I) kynurenic acid, (J) 5-hydroxyindole acetic acid.

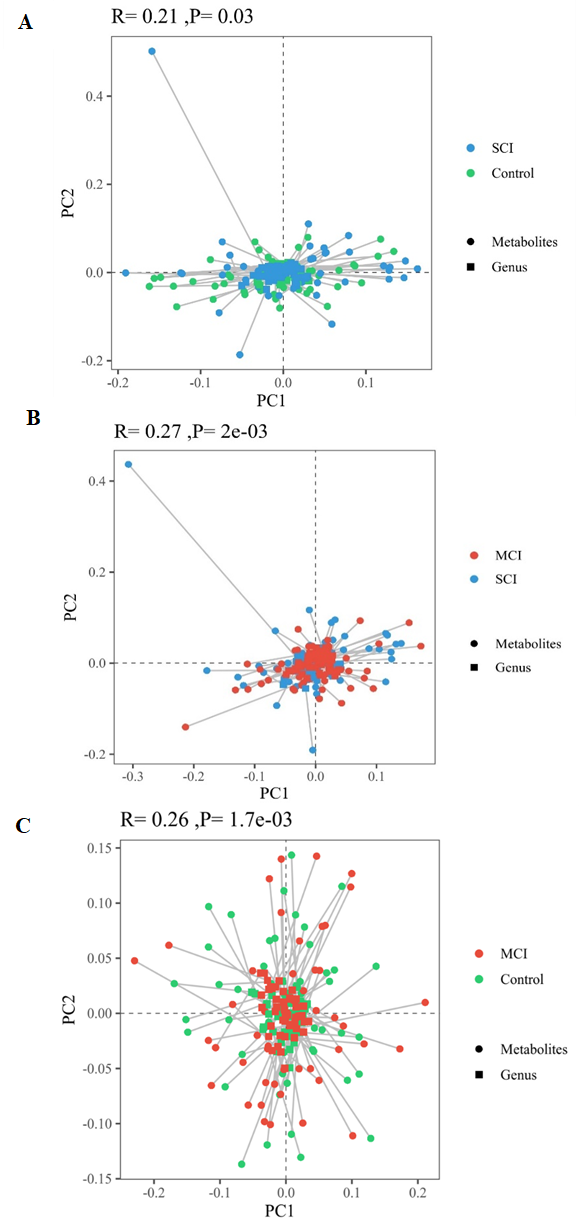

**Figure S3: Serum metabolites and microbiome composition are significantly linked**. Procrustes plot comparing the relationship between the microbiome and the metabolome profiles in control and SCI (A), SCI and MCI (B) and MCI and control (C). Longer lines indicate more within-subject dissimilarity.

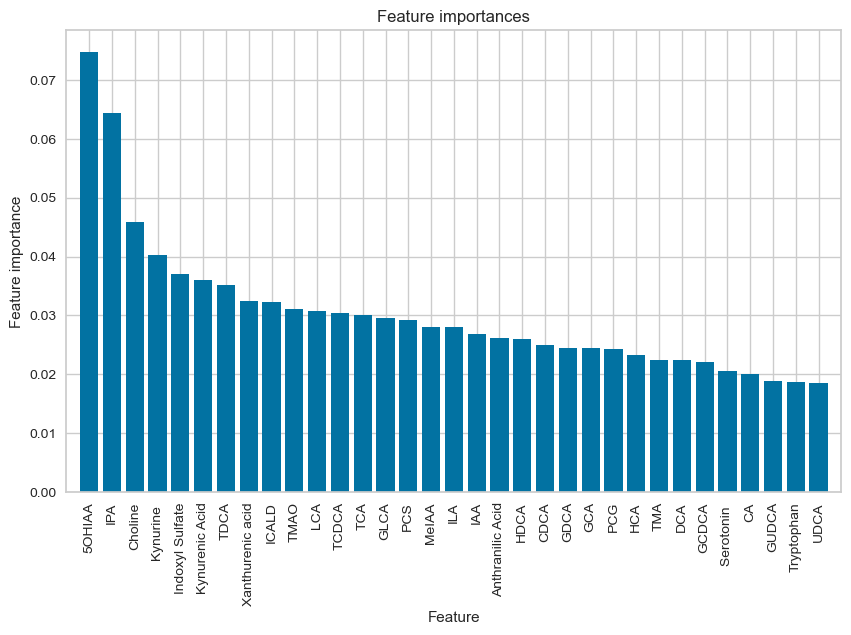

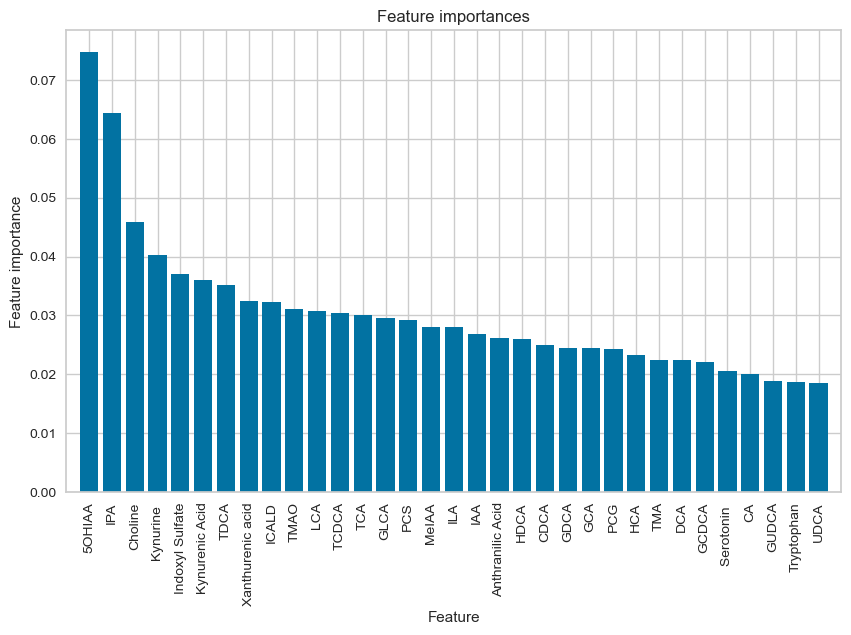

**Figure S4: Identification of the top metabolites predictive of early cognitive decline**. Mean decrease Gini values highlighting the importance of each metabolite in our model. Red box indicates the top six metabolites that gave the highest AUC scores.

| **Supplementary Tables**  **Table S1: Serum metabolite concentrations in control, subjective cognitive impairment (SCI) and mild cognitive impairment (MCI) by LC-MS/MS.** Mean ±SD. P-value generated from one-way ANOVA. <LOD= below the limit of detection. | | | | | |
| --- | --- | --- | --- | --- | --- |
| **Pathway** | **Metabolite** | **Control**  **(µM)** | **SCI**  **(µM)** | **MCI**  **(µM)** | **P-value** |
| TMAO  Pathway | Trimethylamine | 0.94 ± 0.31 | 0.96 ± 0.78 | 1.03 ±1.15 | 0.86 |
|  | Trimethylamine N-oxide | 4.49 ± 2.96 | 5.81± 8.46 | 7.04 ± 9.63 | 0.25 |
|  | Choline | 23.25 ± 6.20 | 19.79 ± 4.93 | 20.09 ± 4.81 | **<0.01** |
| Tryptophan Pathway | Tryptophan | 36.09 ± 4.78 | 36.62 ± 4.44 | 36.08 ± 6.96 | 0.79 |
|  | Kynurenine | 1.07 ± 0.27 | 1.16 ± 0.29 | 1.16 ± 0.27 | 0.19 |
|  | Serotonin | 0.35 ± 0.16 | 0.31 ± 0.17 | 0.35 ± 0.14 | 0.32 |
|  | 5-hydroxyindole acetic acid | 0.05 ± 0.02 | 0.04 ± 0.01 | 0.03 ± 0.01 | **<0.01** |
|  | Kynurenic acid | 0.03 ± 0.01 | 0.04 ± 0.01 | 0.04 ± 0.01 | 0.18 |
|  | Xanthurenic acid | 0.01 ± 0.01 | 0.02 ± 0.01 | 0.02 ± 0.01 | 0.88 |
|  | 3-hydroxyanthranilic acid | <LOD | <LOD | <LOD | - |
|  | Anthranilic acid | 0.07 ± 0.06 | 0.09 ± 0.07 | 0.06 ± 0.06 | 0.11 |
|  | Indole | <LOD | <LOD | <LOD | - |
|  | Indole-3 -acetic acid | 1.33 ± 0.48 | 1.4 ± 0.48 | 1.36 ± 0.68 | 0.84 |
|  | Indole-3- lactic acid | 0.54 ± 0.23 | 0.49 ± 0.14 | 0.50 ± 0.13 | 0.42 |
|  | Indole-3- carboxaldehyde | 0.03 ± 0.01 | 0.03 ± 0.01 | 0.03 ± 0.01 | 0.50 |
|  | Indole-3 -propionic acid | 1.34 ± 0.67 | 1.19 ± 0.62 | 0.96 ± 0.68 | **0.02** |
|  | Methyl indole 3- acetate | 0.03 ± 0.02 | 0.03 ± 0.02 | 0.03 ± 0.02 | 0.41 |
|  | Indoxyl sulfate | 2.41 ± 1.24 | 3.54 ± 2.03 | 3.18 ± 1.55 | **<0.01** |
| *P*-Cresol  Pathway | *P*-cresol sulfate | 19.93 ± 10.87 | 22.79 ± 13.22 | 23.45 ± 12.21 | 0.31 |
|  | *P*-cresol glucuronide | 0.13 ± 0.14 | 0.13 ± 0.20 | 0.12 ± 0.12 | 0.87 |
| Bile Acid  Pathway | CA | 0.23 ± 0.35 | 0.26 ± 0.48 | 0.24 ± 0.40 | 0.95 |
|  | CDCA | 0.01 ± 0.01 | 0.01 ± 0.02 | 0.01 ± 0.02 | 0.71 |
|  | HDCA | 0.02 ± 0.02 | 0.02 ± 0.02 | 0.03 ± 0.06 | 0.17 |
|  | GCDCA | 0.39 ± 0.26 | 0.49 ± 0.90 | 0.41 ± 0.33 | 0.64 |
|  | GDCA | 0.16 ± 0.15 | 0.20 ± 0.33 | 0.23 ± 0.24 | 0.46 |
|  | GCA | 0.10 ± 0.08 | 0.23 ± 0.84 | 0.14 ± 0.19 | 0.43 |
|  | DCA | 0.26 ± 0.26 | 0.27 ± 0.31 | 0.31 ± 0.45 | 0.75 |
|  | TCDCA | 0.05 ± 0.05 | 0.10 ± 0.35 | 0.06 ± 0.07 | 0.43 |
|  | TDCA | 0.02 ± 0.02 | 0.04 ± 0.10 | 0.05 ± 0.07 | 0.21 |
|  | GUCDA | 0.06 ± 0.08 | 0.05 ± 0.07 | 0.05 ± 0.06 | 0.75 |
|  | TUDCA | <LOD | <LOD | <LOD | - |
|  | TLCA | <LOD | <LOD | <LOD | - |
|  | THCA | <LOD | <LOD | <LOD | - |
|  | GLCA | 0.02 ± 0.02 | 0.01 ± 0.01 | 0.03 ± 0.06 | 0.19 |
|  | GHCA | <LOD | <LOD | <LOD | - |
|  | GHDCA | <LOD | <LOD | <LOD | - |
|  | UDCA | 0.03 ± 0.05 | 0.04 ± 0.07 | 0.03 ± 0.06 | 0.83 |
|  | TCA | 0.03 ± 0.03 | 0.09 ± 0.40 | 0.04 ± 0.06 | 0.41 |
|  | HCA | 0.01 ± 0.01 | 0.01 ± 0.01 | 0.01 ± 0.1 | 0.39 |
|  | LCA | 0.03 ± 0.03 | 0.04 ± 0.03 | 0.05 ± 0.07 | 0.13 |

| **Table S2: Comparison of area under the receiver operator curve (AUC)** | | | |
| --- | --- | --- | --- |
| Machine Learning Model | Classification | | |
|  | Control vs. SCI vs. MCI  (32 metabolites)  AUC | Control vs. SCI vs. MCI  (6 metabolites)  AUC | Control vs. MCI  (6 metabolites)  AUC |
| Random Forest | 0.65 | 0.74 | 0.84 |
| AdaBoost | 0.58 | 0.68 | 0.87 |
| Naïve Bayes | 0.63 | 0.72 | 0.90 |
